# Sanitation and Diarrheal Morbidity: Evidence from Afghanistan

**DOI:** 10.1101/2020.10.20.20216333

**Authors:** Manzoor Ahmad Malik, Saddaf Naaz Akhtar

## Abstract

**Introduction:** Lack of sanitation facilities and inadequate water is key to many diseases’ outcomes, mainly diarrhea. These factors not just affect the health status of a country and but also result in increased mortality and morbidity pattern, particularly among the vulnerable population like children and women. Thus, the study assesses the linkages between diarrheal diseases and sanitation in Afghanistan affected since long by political instability which has derailed the health system of the country.

**Material and Methods:** We used 2015 Afghanistan Demographic and Health Survey to assess the linkages between sanitation and diarrheal diseases among under-five children. Bivariate and multivariate analyses have used to carry out the study. The logistic model was used to evaluate the risk factors that lead to diarrheal outcomes.

**Results:** Our findings from the results showed that the prevalence of diarrhea among under-five children in Afghanistan was 29%. Pashai is the most affected, with 36% among ethnicities, whereas eastern Afghanistan is the most affected region with the prevalence of (38%). Open defecated Population (OR: 1.17, p<0.001), Tap water (OR: 1.31, p<0.001), Well water (OR: 1.24, p<0.001), and Sharing toilet (OR: 1.15, p<0.001) are significantly associated with childhood diarrhea.

**Conclusions:** A significant impact has found with factors like-inadequate sanitation, shared toilet facility, clean water and other elements with childhood diarrhea in Afghanistan. The region-wise difference has also found to be very high across the regions. Thus, it has found that a lack of such factors has a more significant impact on the health of children and needs a particular focus from a policy purpose.

## Introduction

Morbidity patterns resulting due to pneumonia, diarrhea, fever, and acute respiratory infection are vital in increasing the risk of mortality among the under-five children [1,2]. Diarrheal diseases constitute a significant problem among under-five children across the globe after pneumonia [3,4]. Children in the developing world are positively affected by diarrheal diseases, which can be preventable and eradicated by possible interventions [5,6]. Previous studies have shown that children in developing countries are ten times more likely to die before the age of 5 years than children in the developing world [7,8]. Empirical studies reflect this fact that one-fourth of these deaths among children result in south Asia among the under-five children and Afghanistan is one of the most affected countries in the region [9– 12,12,13]. According to the 2015 Afghanistan Demographic and Health Survey (AfDHS) report, around one in every 18 children die before completing the age five in Afghanistan, and these deaths mostly occur in the very first year of birth among children [14]. The risk factors to these deaths are manifold and result from both demand and supply-side factors. Other things include the lack of spending in the health care system and socio-behavioral and contextual factors.

Diarrhea is the second leading cause of deaths, accounting nearly one in every nine deaths worldwide [15]. It is one of the largest reasons for diseases burden in Afghanistan after pneumonia [16].

The above figure shows the percentage of deaths due to diarrhea in Afghanistan since 2000 based on UNICEF data and the results show that although there is a decline in diarrheal deaths, still it accounts for nearly 9% of deaths. A recent survey of 2015 AfDHS showed that around 29% of children have an acute diarrheal disease in Afghanistan. Therefore, the present study tries to explore the possible factors that affect the greater prevalence of diarrhea among under-five children based on the recent round of 2015 AfDHS.

### Theoretical Context

According to UNICEF, around 2.4 billion people lack adequate sanitation facilities across the world, and approximately 663 million people don’t have access to improved water [5]. However, the SDG have aimed at improving the conditions of health, particularly of children and women. But these challenges persist and affect at a grander scale through morbidity and mortality patterns among under-five children, particularly in developing countries. Children at younger ages are at greater risk to water-borne diseases like diarrhea, which affect their health and wellbeing and put them at greater risk. Around 800,000 children die yearly due to conditions resulting through lack of sanitation facilities which can be cured [15]. Diarrhea has become one of the leading causes of morbidity and mortality among children [17]. Though the diseases have attributed to many factors but some closely associated reasons are inadequate water and lack of access to sanitation facilities [18]. These factors affect the burden of diarrheal deaths and hence contribute to a greater prevalence of acute diarrheal diseases [19]. While the socio-demographic factors have played an important role, but factors like-open defecation, unhygienic practices, unimproved water, etc. have shown a significant effect [20]. Numerous studies in this context have found that households having inadequate sanitation facilities have a more substantial impact on the incidence of diarrheal outcomes [21,22]. A study [23] showed that the availability of drinking water is an essential factor and lack of it results in a greater incidence of childhood diarrheal deaths. A report on Sustainable Development Goals in 2017 also reflected the fact that higher risk for infectious diseases like diarrhea is mainly due to the lack of safe water sanitation and other hygiene services [24]. Increased risk of diarrhea is also due to the proximity of sanitation facilities to homes, sharing of sanitation facilities, and poor hygiene [25].

Diarrheal deaths account nearly 1.87 million deaths annually resulting due to numerous factors but primarily due to unsafe water and inadequate sanitation [7,9]. Better sanitation facilities are essential to reduce the risk of diarrheal morbidity [26]. Studies clearly show that sanitation infrastructure is most effective, reducing diarrhea incidence by about 20% while as clean water by 11% [27]. It has observed that interventions in providing better access to clean water and toilet facilities lower the risk of diarrhea reduction ranging from 27% to 53% among the children aged below five years [28]. Improved sanitation and hygiene are essential to avert the impact of diarrheal deaths [29]. But a better standard of living conditions can also help to reduce the burden of diarrhea among under-five children [30]. Thus, targeting the measures which can reduce the levels of inadequate sanitation and lack of access to clean water can help to lower the morbidity patterns resulting due to diarrhea among children aged below five particularly in a developing world [31].

Furthermore, hygiene improvement effectively reduces diarrhea and is a critical element of child health and nutritional promotion [32–34]. Though the evidence is strong despite this, there has been little sound evidence published so far on to what extent the availability and utilization of latrines and better water facilities can reduce diarrheal prevalence [35,36] and the country like Afghanistan has least explored. Therefore, this study tried to examine the linkages between diarrhea and its possible association with sanitation and inadequate water facilities.

### Conceptual Definitions

Diarrhea is defined as the passage of three or more liquid stools within 24 hours [26]. Diarrhea can last for several days, but According to this study, acute diarrhea is the condition where a child suffers from diarrhea for less than 14 days since this leads to severe dehydration and loss of fluids resulting in diarrheal deaths. According to WHO other causes like septic bacterial infections also account for more diarrheal deaths. Whereas medically it is defined as a symptom of infection in the intestinal tract, resulting due to bacterial, viral and parasitic organisms resulting through unimproved water, inadequate sanitation and poor hygiene.

Sanitation is critical to promote not just human health but also have socio-demographic conditions like food security, women empowerment, girl education, social security, and reduction in loss of morbidity and mortality. It just not reduces intestinal and vector-borne diseases but also has a significant impact on diarrhea [28]. Sanitation, in simple words, means the provision of facilities and services for the safe disposal of human urine and faeces. Sanitation refers to the maintenance of hygienic conditions, through services such as garbage collection and wastewater disposal. The study uses various measures to measure the sanitation access, such as lack of adequate water, toilet facility, access to water and toilet sharing facility.

## Material and Methods

The data in this study has taken from Afghanistan Demographic and Health Survey (AfDHS) conducted in 2015-16. AfDHS surveys consist of data on a wide range of public health topics, including anthropometric, demographic, socioeconomic, family planning, and domestic violence issues. The AfDHS (2015-16) provides up-to-date information on the socio-demographic characteristics of the respondents between the ages of 15–49 from randomly selected households. AfDHS is a national sample survey that provides up-to-date information on fertility levels; marriage; fertility preferences; awareness and use of family planning methods; child feeding practices; nutrition, adult, and childhood mortality; awareness and attitudes regarding HIV/AIDS; women’s empowerment; and domestic violence.

### Study Participants

The total sample for this analysis was 30303 children, aged 0_59 months, who had complete morbidity data and were living with their mothers at the time of the survey. The AfDHS collected data on morbidity such as diarrhea which is defined as the passing of three or more liquid, watery or loose stools per day. The data was collected based on a survey question to mothers, whether any of their children below five years of age had diarrhea during the preceding weeks in the survey.

The outcome variable was the prevalence of diarrhea during the two weeks. This question was asked to Mothers, whether or not their child suffered from diarrhea during the past two weeks. The leading independent variables are proxies of indicators like sanitation facility, drinking water, apart from other socio-demographic variables like age of the child, sex residence, and region.

Bivariate and Multivariate analysis was carried out in the paper to study the association between sanitation and diarrhea. We used a logistic model with a dependent variable categorized into a binary outcome variable as diarrhea and regress it with other closely associated risk factors.

## Results

Figure 2 shows the prevalence of diarrhea by regions. Diarrheal prevalence has found to be higher in the Eastern region (39%) of Afghanistan followed by West (31%) and North (30%) regions. In contrast, lower prevalence has found to be in the Sothern region of Afghanistan (23%).

**Fig. 1:**
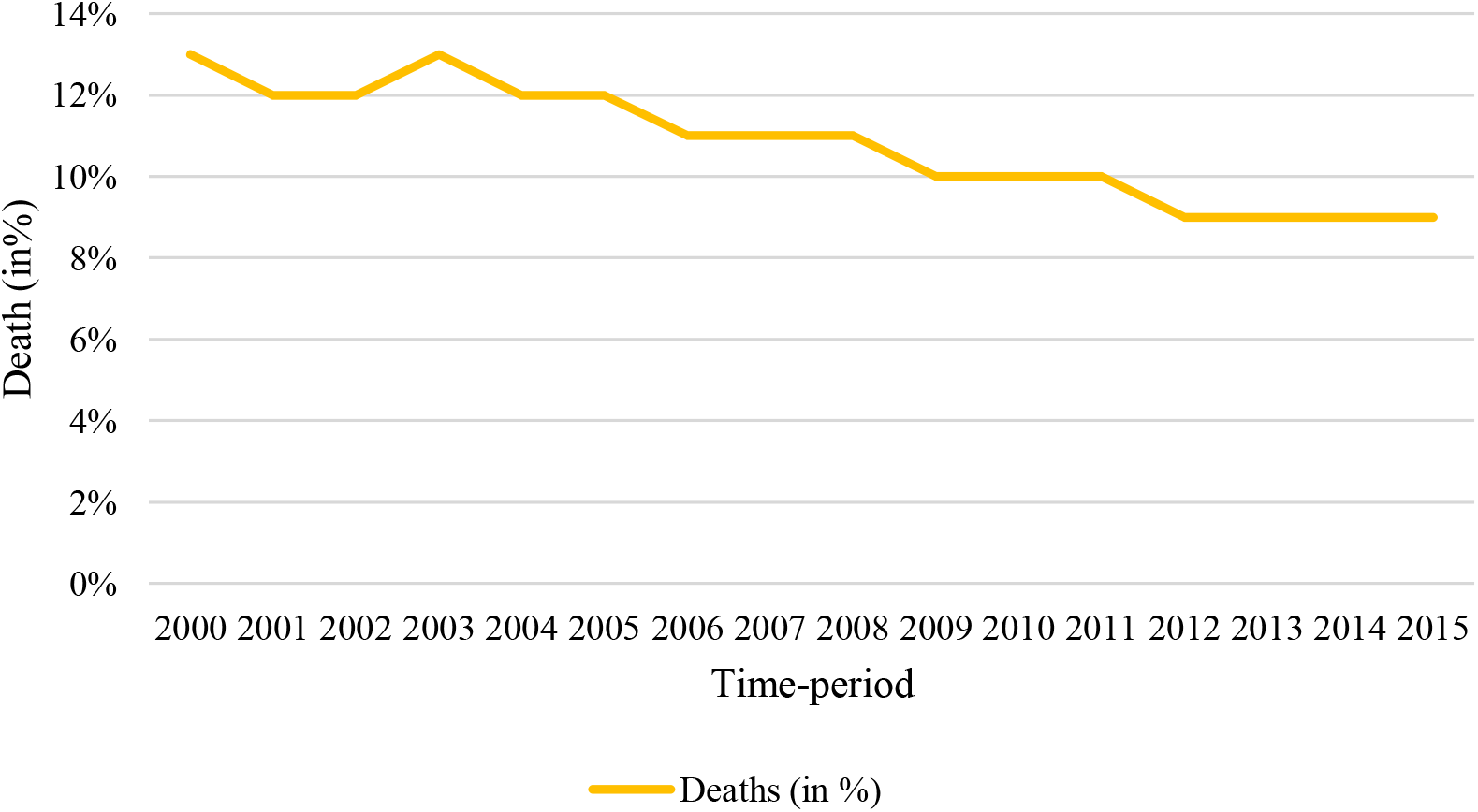
Percentage of deaths due to diarrhea in Afghanistan from 2000-2015. Source: UNICEF

**Fig. 2:**
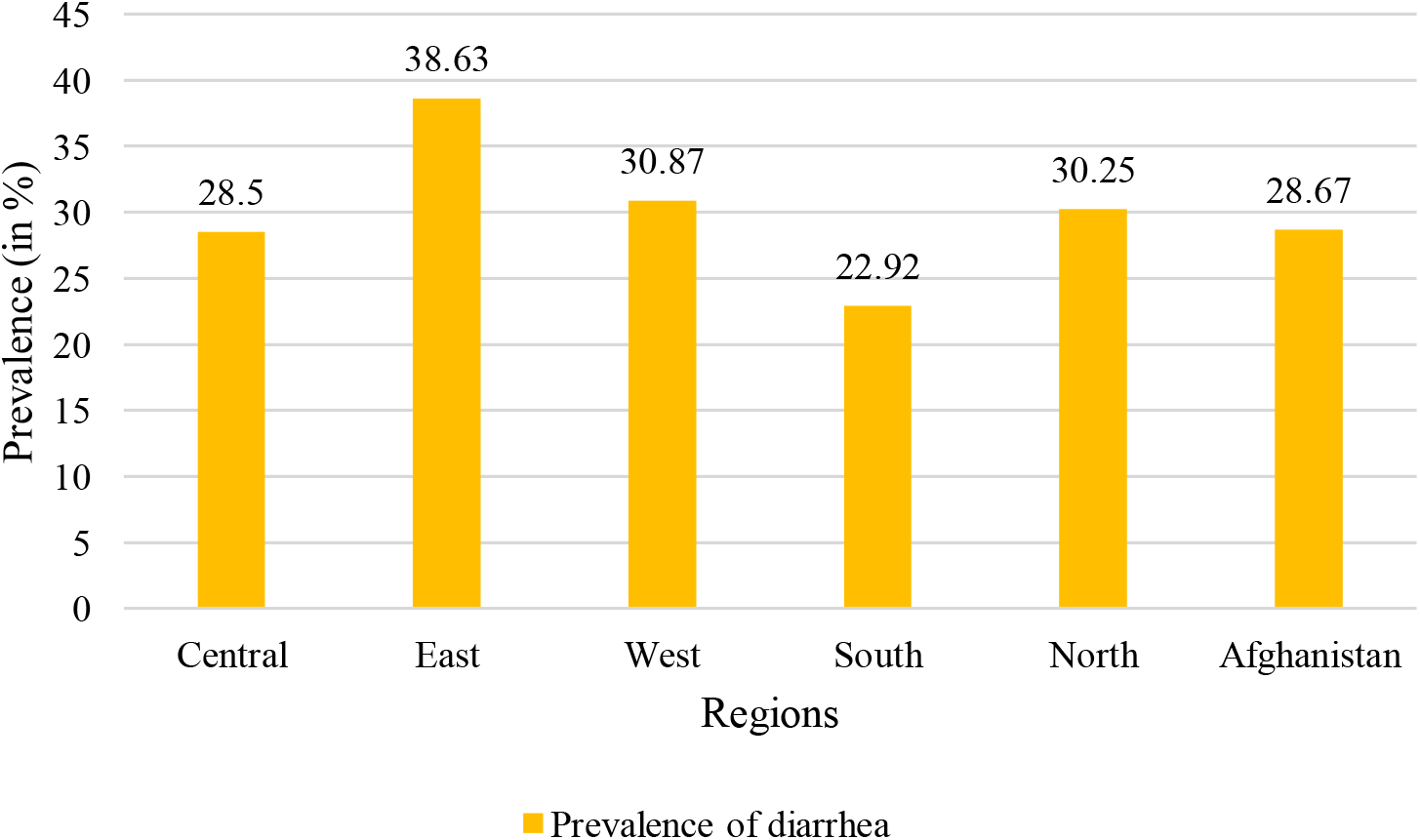
Prevalence of diarrhea by regions in Afghanistan in 2015. Source: AFDHS-2015

Figure 3 shows the prevalence of diarrhea by ethnic groups which has found to be highest among the Pashai (35%) followed by Uzbeks (33%). The lowest prevalence, although was found to be among the Balochs, which was just at 15 %.

**Fig. 3:**
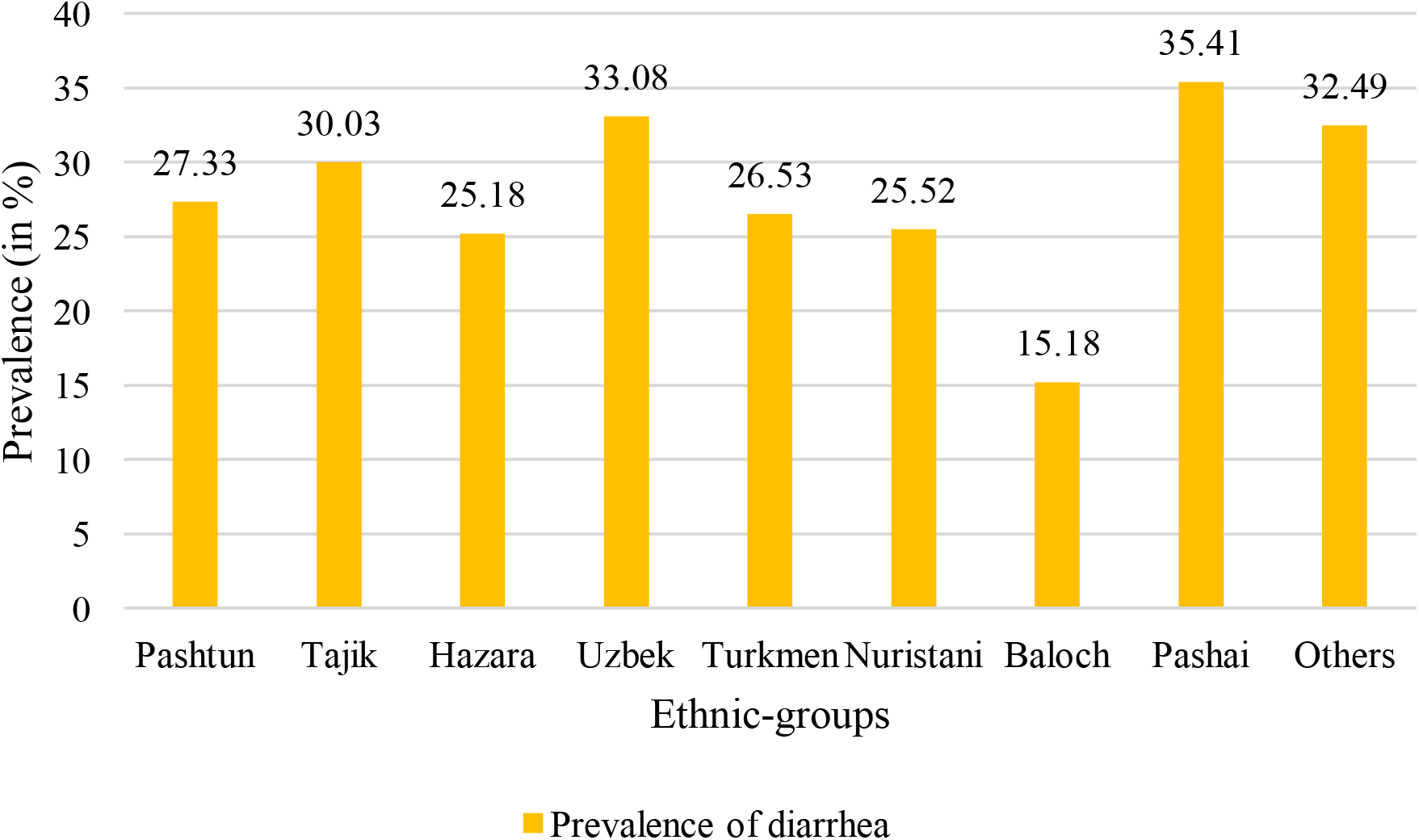
Prevalence of diarrhea by ethnic-groups in Afghanistan in the year 2015. Source: AFDHS-2015

Table 1 shows the prevalence of diarrhea along with according to demographic and socio-economic characteristics prevalence of diarrhea has found to be higher among the children aged 6-12 and 13 to 24 months, similarly, we see prevalence higher among female children to that of male children. We found diarrhea among children was found to be higher among working women. In contrast, children of adolescent mothers are having a 2% higher prevalence of diarrhea than the children of adult mothers. Households defecating in the open are having a higher prevalence of diarrhea among children. Similarly, it was to found to be higher among those households sharing toilets. Similar things have found to be among the other water and sanitation factors as can be seen in the table.

**Table 1.** Prevalence of Diarrhea with Background Characteristics AfDHS-2015.

| Background characteristics | Prevalence of Diarrhea (%) | Sample distribution |
| --- | --- | --- |
| ***Child age*** |  |  |
| <6 | 20.83 | 3095 |
| 6-12' | 34.61 | 2720 |
| 13-24' | 37.92 | 5708 |
| 25-36' | 32.16 | 6598 |
| 49+ | 19.11 | 5902 |
| ***Child sex*** |  |  |
| Male | 29.49 | 15605 |
| Female | 27.8 | 14699 |
| ***Birth order*** |  |  |
| <2 | 28.85 | 11159 |
| 3-5' | 27.86 | 11640 |
| 6+ | 29.64 | 7505 |
| ***Mother's education*** |  |  |
| Education | 28.47 | 25261 |
| Primary | 32.1 | 2429 |
| Secondary | 29.6 | 2130 |
| Higher | 17.77 | 484 |
| **Mother's working status** |  |  |
| No | 28.28 | 26925 |
| Yes | 32.31 | 3244 |
| **Mother's age at first birth** |  |  |
| Adolescent Mothers | 29.67 | 17612 |
| Adult Mothers | 27.27 | 12692 |
| **Place of residence** |  |  |
| Urban | 32.01 | 7040 |
| Rural | 27.65 | 23264 |
| **Sources of drinking water** |  |  |
| Piped water | 25.34 | 1898 |
| Tap water | 30.61 | 4918 |
| Well water | 29.05 | 13716 |
| Unimproved water | 27.8 | 9771 |
| **Time taken to Water** |  |  |
| Premises | 27.64 | 13625 |
| Out of Premises | 29.68 | 16393 |
| **Toilet facility** |  |  |
| Open Defecated | 30.1 | 4016 |
| Non-Open Defecated | 28.45 | 26288 |
| **Toilet sharing facility** |  |  |
| No | 28.08 | 25416 |
| Yes | 31.73 | 4888 |
| **Breast feeding** |  |  |
| Never breast feeding | 28.67 | 507 |
| Ever breast feeding | 28.6 | 29797 |
| **Regions** |  |  |
| Central | 28.50 | 6021 |
| East | 38.63 | 1742 |
| West | 30.87 | 4426 |
| South | 22.92 | 7470 |
| North | 30.25 | 10646 |
| Afghanistan | 28.67 | 30304 |

**Sources:** Authors' calculation from AfDHS-2015.

Table 2 studies the association of diarrhea along with various determinants of sanitation and water and some vital demographic factors. We found that age is closely associated with diarrhea, particularly from 6 to 18 months (OR= 2.26 & 2.45; p<0.001). Similarly, female children are 6% less likely to have diarrhea as compared to that of males (OR= 0.94; p<0.001). Our results showed that children belonging to households defecating in the open are 17 times more likely to have diarrheal diseases as compared to those which don’t (OR= 1.17; p<0.001). Similarly, we can see that children belonging to households sharing toilets are at greater risk of diarrheal diseases (OR= 1.15; p<0.001). At the same time, it has found that those using tap water are at greater risk of having diarrheal diseases followed by well water (OR= 1.31 & 1.24 p<0.001). Similarly, those taking another form of water are also at a greater risk of having any diarrheal diseases with an odds ratio of 1.12 at 10% significance level.

**Table 2.** Association of diarrhea with key contextual factors in Afghanistan-2015.

| Background characteristics | Odds Ratio | Confidence Interval |  |
| --- | --- | --- | --- |
|  |  | Lower limit | Upper limit |
| **Child age** |  |  |  |
| <6® | 2.26*** | 2.00 | 2.56 |
| 6-12' | 2.45*** | 2.20 | 2.73 |
| 13-24' | 2.05*** | 1.85 | 2.29 |
| 25-36' | 1.61*** | 1.45 | 1.80 |
| 49+ | 1.09 | 0.97 | 1.22 |
| **Child sex** |  |  |  |
| Male® |  |  |  |
| Female | 0.94** | 0.89 | 0.99 |
| **Birth order** |  |  |  |
| <2® |  |  |  |
| 3-5' | 1.03 | 0.97 | 1.10 |
| 6+ | 1.06* | 1.00 | 1.14 |
| **Place of residence** |  |  |  |
| Rural® |  |  |  |
| Urban | 0.86*** | 0.81 | 0.93 |
| **Sources of drinking water** |  |  |  |
| Piped water® |  |  |  |
| Tap water | 1.31*** | 1.14 | 1.52 |
| Well water | 1.24*** | 1.09 | 1.42 |
| Unimproved water | 1.12* | 0.98 | 1.29 |
| **Time taken to water** |  |  |  |
| Premises® |  |  |  |
| Out of Premises | 1.05 | 0.99 | 1.11 |
| **Toilet facility** |  |  |  |
| Open Defecated® |  |  |  |
| Non-Open Defecated | 1.17*** | 1.09 | 1.27 |
| **Toilet sharing facility** |  |  |  |
| No® |  |  |  |
| Yes | 1.15*** | 1.07 | 1.25 |
| **Regions** |  |  |  |
| Central® |  |  |  |
| East | 1.72*** | 1.55 | 1.92 |
| West | 1.09* | 1.00 | 1.20 |
| South | 0.58*** | 0.54 | 0.63 |
| North | 0.96 | 0.90 | 1.04 |
| **Notes:** ®: reference category; *p<0.1; ** p<0.05; *** p<0.01; **Sources:** Authors' calculation from AfDHS-2015. |  |  |  |

## Discussion

Child health is an important issue to be addressed around the globe. Deteriorating child health also have long-run consequences on the health system of a country apart from its socio-economic adversities. Child health is affected by various diseases ranging from diarrhea to acute respiratory infections due to vulnerability and less immune systems of children at early ages than the other population groups [37]. Furthermore, the lack of better access to sanitation and clean water results in the greater vulnerability of children at lower age and put them at higher risk. It has found that nearly 90% of diarrheal deaths occur among children in developing countries [38–40]. Studies show that low-income countries are not just affected by deficient water system but for higher levels of open defecation and lack access to adequate drinking water [7,41].

Afghanistan is one of the most affected regions in the world due to conflict, and the health system is one of the poorest. While it is evident that higher rates of diarrhea prevail due to multiple risk factors ranging from socio-demographic to economic factor [42,43]. But our study focused mainly on the aspects related to sanitation and water-hygiene. While examining a few behavioral factors like age, sex of the selected population, we found their impact very much consistent with the earlier studies [43–47]. To address the key purpose of this paper, we studied the factors like open defecation, shared toilet facility, access to drinking water and drinking water sources in Afghanistan. The results showed a significant association between diarrhea with these essential factors. It has found that piped water supplied to households was a key risk factor with greater odds of affecting the diarrheal diseases in Afghanistan. Studies have clearly shown that the sanitation facility has correlated with diarrheal morbidity, incredibly open defecation [48–53]. While examining the linkages between open defecation and diarrhea our result was consistent with the earlier findings.

Various factors are related to diarrheal diseases, particularly poor domestic sanitation water quality, service level and hygiene [30]. Afghanistan is one of the worlds affected regions in the world, both in terms of health standards of living. It is a country having the highest IMR poverty rates and other socio-demographic indicators [54,55]. Diarrheal disease still claims the lives of 26 children each day across the country, accounting nearly 12% of deaths in Afghanistan [56]. So, understanding these contextual factors associated with diarrheal diseases in the country are key. These identified factors have a significant impact due to externality effects and hence need a particular focus, particularly at the regional level as indicated by our study. Further studies can be conducted to estimate the burden of these factors on morbidity pattern. Since it is already clear from the above that poor sanitation like open defecation, shared toilet facility and lack of access to drinking water are leaving children more susceptible to infections that cause diarrhea.

## Conclusions

The study tried to examine the linkages between sanitation and diarrhea in Afghanistan based on the DHS survey conducted in 2015. Our study is consistent with many other studies while examining the associations between open defecation and diarrheal diseases. We also found that a lack of water and toilet have a significant impact on the health of children. Diarrhea has found to be closely associated with lack of access to proper drinking water and inadequate sanitation facility. Thus, to reduce the burden of diarrheal diseases, it is essential to focus on the sanitation facilities apart from providing access to clean and purified water. Inclusive policy approach can enhance the health challenges and also give positive externalities in terms of public health challenges. Afghanistan needs to focus on the bettering the sanitation and water hygiene facilities so that the more significant burden of diarrheal diseases can be averted to improve the child health.

## Data Availability

The data in the present study has taken from Afghanistan Demographic and Health Survey (AfDHS) conducted in 2015-16. This is online available.

https://dhsprogram.com/what-we-do/survey/survey-display-471.cfm

## Acknowledgement

None.

